# Conductive Hydrogels as a Medium for Shapeable Catheter Ablation Electrodes

**DOI:** 10.64898/2026.10.01.26364546

**Authors:** Drew Bernard, Lukas Jaworski, Mathews John, Allison Post, Abbey Nkansah, Derek Bashe, Meng-Hsuan Sung, Elizabeth Cosgriff-Hernandez, Mehdi Razavi

**Author notes:** Correspondence to: Mehdi Razavi, MD, Division of Cardiology, Department of Internal Medicine, Baylor College of Medicine, 6624 Fannin, #2480, Houston, Texas 77030, USA. These authors contributed equally to this work (Drew Bernard, Lukas Jaworski).

## Abstract

**BACKGROUND:** Radiofrequency catheter ablation (RFCA) uses rigid metallic electrodes whose fixed geometry limits tissue contact, lesion continuity, and safe energy delivery near vulnerable structures. We evaluated a moldable, tissue-conforming conductive hydrogel as an RFCA electrode *ex vivo*.

**METHODS:** Poly(ether urethane diacrylamide) hydrogels with saline-tuned conductivity were molded into plugs, disks, and bars and affixed to an irrigated 8-Fr catheter. Ablations were performed on excised porcine tissue in circulated saline at 37 °C under fixed contact force and irrigation. Lesion geometry, intramural temperatures, and steam pops were compared with bare-metal (BM) electrodes and size-matched metallic analogs; an atrioesophageal stack model assessed collateral injury. Lesions were delineated using triphenyltetrazolium chloride staining.

**RESULTS:** Hydrogel bars produced wide, ellipsoidal lesions (width 10.51 ± 1.40 versus 8.71 ± 1.65 mm for BM; *P*=0.031; aspect ratio 1.72); size-matched metal bars produced none. Hydrogel disks produced lesions statistically indistinguishable from BM in depth (4.54 ± 0.90 versus 4.84 ± 0.55 mm) and width (9.47 ± 0.89 versus 11.41 ± 2.39 mm), whereas metal disks produced significantly shallower lesions (2.30 ± 1.70 mm; *P*=0.015 versus BM). In the atrioesophageal model, hydrogel plugs achieved transmural atrial lesions with a reduced esophageal lumen temperature (56.33 ± 3.80 versus 61.30 ± 5.14 °C; *P*=0.046), total lesion depth (4.03 ± 1.25 versus 7.58 ± 0.78 mm; *P*<0.001), and esophageal lesion width (3.05 ± 2.35 versus 8.70 ± 2.69 mm; *P*<0.001) compared with BM. Hydrogel electrodes reduced steam-pop risk (relative risk 0.34; 95% CI 0.13–0.88). Plug characterization confirmed saline-dependent conductivity and attenuated energy delivery, with lower subsurface peak temperatures and shallower lesions than BM.

**CONCLUSIONS:** Moldable, conductivity-tunable hydrogels shape RF lesions and may improve ablation safety, supporting evaluation for contiguous or linear lesion sets and ablation near sensitive structures.

**CLINICAL PERSPECTIVE:** *What Is Known:* • Radiofrequency ablation catheters use rigid metallic electrodes, so contiguous lesion sets require sequences of closely spaced burns, and irregular tissue surfaces or thin atrial tissue near the esophagus limit uniform, safe energy delivery. • Conductive hydrogels can be molded into arbitrary shapes and tuned in electrical conductivity, but their use as a radiofrequency ablation electrode medium has not been characterized.

*What the Study Adds:*

- In an ex vivo porcine model, a molded hydrogel bar electrode produced wide, ellipsoidal lesions where a size-matched metal bar produced none, and a hydrogel disk produced lesions comparable to a standard bare-metal electrode where the size-matched metal disk was significantly shallower.
- Hydrogel electrodes attenuated energy delivery, achieving transmural atrial lesions with lower esophageal lumen temperatures, smaller esophageal lesions, and lower steam-pop risk than a bare-metal electrode.
- These properties support conductive hydrogel electrodes for linear lesion sets and ablation near sensitive structures, pending in vivo validation.

## INTRODUCTION

Treatments for arrhythmias include antiarrhythmic medications and interventions such as radiofrequency catheter ablation (RFCA) and pulsed field ablation to control fibrillatory circuits. Antiarrhythmic drug therapy alone successfully resets rhythm in only approximately 57% of patients, and under some conditions these drugs can be proarrhythmic.^1^ Likewise, after cardiac ablation, reentrant circuit reconnections can form from insufficient lesions around the arrhythmia substrate; thus, although cardiac ablation is typically more effective than antiarrhythmic medication management only, it is associated with a recurrence rate as high as 45%.^2^ On the other hand, pulsed field ablation has been shown to be safe and effective for treating atrial arrhythmias, but it cannot create the deeper lesions required to treat ventricular arrhythmias originating beneath the endocardial surface.

RFCA catheters, which are the current clinical standard, operate by delivering a controlled high-frequency electrical current into tissue to create focused thermal damage around the point of contact. Typical ablation catheter designs include one or more rigid metallic electrodes that transmit energy into tissue; these electrodes limit the application of radiofrequency energy to areas with sufficient electrode contact. Thus, forming a contiguous lesion set requires performing a sequence of closely spaced ablations.^3,4^

Some catheters have been designed to overcome this limitation by delivering energy from multiple electrodes with a large effective contact area to create larger lesions.^5–7^ Using metallic electrodes, which are inherently rigid, makes it difficult to apply RFCA in areas where the cardiac tissue surface is uneven. For example, inside the trabeculated pectinate muscles of the atrial appendage or around papillary muscles, it can be difficult to apply ablative energy uniformly to the tissue because the electrode makes only limited contact with it. Rigid electrodes also deliver energy without regard to what lies beyond the target wall, so ablation of thin atrial tissue near the esophagus carries a well-recognized risk of collateral thermal injury.^8^

Conductive hydrogels can be tuned—in both mechanical and electrical properties—to form flexible electrodes that can better conform to the target tissue and transmit RF energy to tissue. In this study, we used an ex vivo model to assess the use of a flexible and conductive hydrogel in RFCA electrodes to avoid the limitations of rigid metallic electrodes. We characterized how the hydrogel’s electrical conductivity and molded shape govern lesion geometry and intramural heating, benchmarking each against equivalently sized rigid metallic electrodes. We then tested the hydrogel in an atrioesophageal model to determine whether its conformability and energy-modulating behavior could achieve transmural atrial lesions while limiting off-target heating of adjacent structures.

## MATERIALS AND METHODS

### Ex Vivo Ablation Setup

The ex vivo model used in this study (Figure 1A, Figure S1) is based on previously published models.^8,9^ Ablations were performed on freshly excised porcine hearts (Animal Technologies, Tyler, TX) with an irrigated ablation catheter connected to either a Stockert 70 RF Generator (Biosense Webster, Irvine, CA) or a Stockert SmartAblate Generator (Biosense

**Figure 1.**
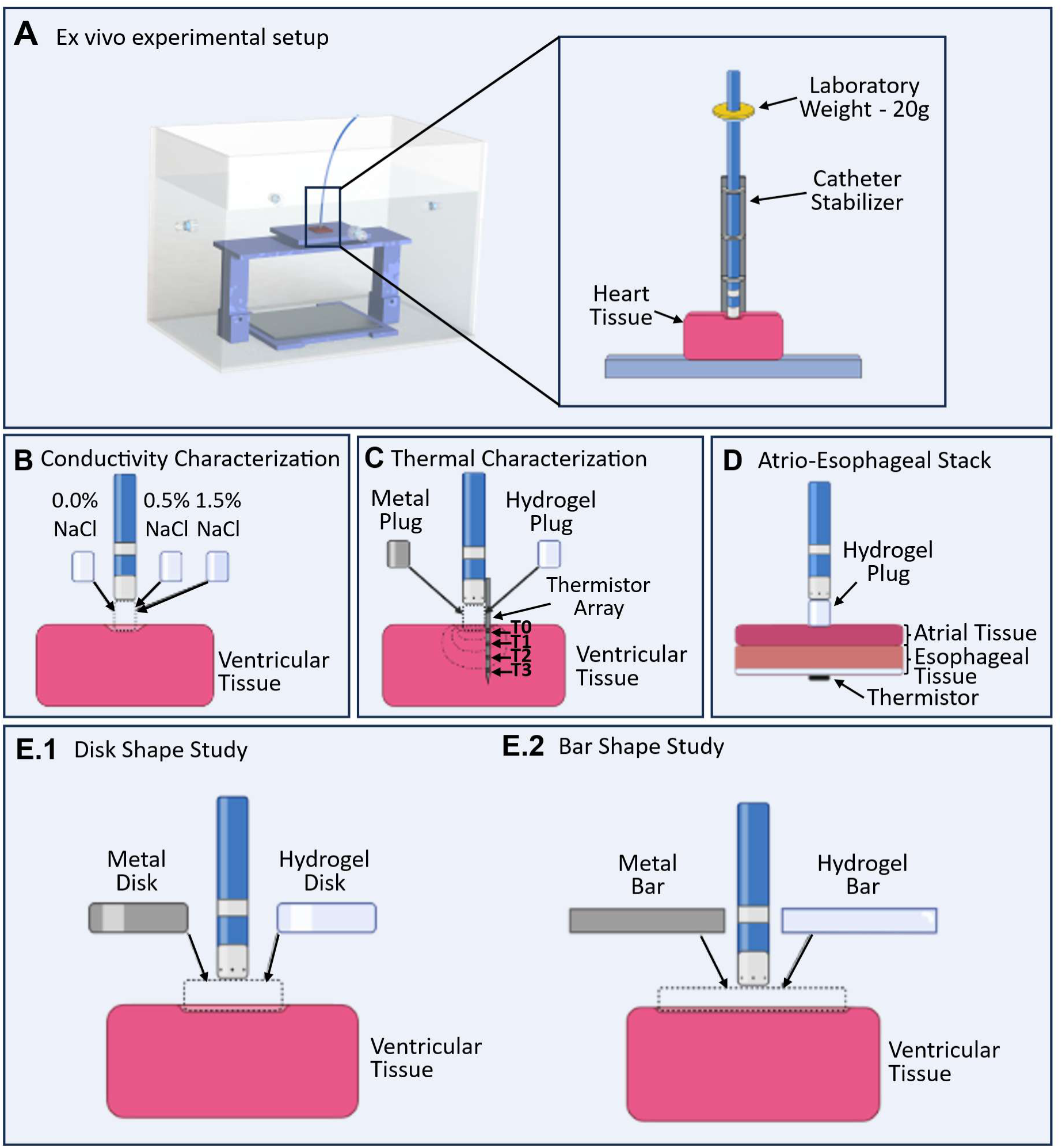
General experimental setup to characterize lesions. (**A**) Rendered view of the tank setup used in each experiment; catheter setup, with a 3D-printed catheter stabilizer and a 20-g laboratory weight to provide a constant downward contact force (inset). (**B**) Electrical conductivity of hydrogel electrodes was tuned by hydrating 2.5 × 3.25-mm hydrogel plug electrodes with deionized water, 0.5% NaCl, or 1.5% NaCl; the resultant lesion geometry was compared with the geometries of lesions formed by a standard ablation catheter. (**C**) Geometries and thermal profiles were compared between lesions formed with a 2.5 × 3.25-mm hydrogel plug electrode and lesions formed by a metallic equivalent and a standard ablation catheter; tissue temperatures were measured from a custom thermistor array inserted into the left ventricular tissue sample. (**D**) Application of 2.5 × 2.0-mm hydrogel plug electrode in atrial ablation; an atrio-esophageal stack was used with a temperature probe placed on the endoluminal side of the esophagus. Conductive hydrogel was shaped into (**E.1**) 10 × 2.0-mm round electrode configurations and (**E.2**) 5 × 30 × 2.5-mm bar electrode configurations; lesion geometries were characterized and compared with those of lesions formed by both a metallic equivalent and a standard ablation catheter electrode.

Webster, Irvine, CA). Because the catheter used is not force-sensing, a constant 20-g load was applied to the catheter by laboratory weights and is referred to hereafter as the applied contact force; irrigation was maintained at 30 mL/min for all ablations. A saline tank maintained at 37 °C was prepared, with a tissue stage inside on which the tissue samples were positioned. The ablation catheter was held perpendicular to the tissue stage. A metal plate positioned underneath the stage formed the grounding pad. No live-animal procedures were performed, so institutional animal care and use committee review was not required. Tissue samples for each experiment were taken from several hearts, and each ablation was treated as an independent observation.

### Hydrogel Preparation

Individual poly(ether urethane diacrylamide) (PEUDAm) hydrogels were prepared according to previously published methods.^10^ The hydrogels were molded into various shapes depending on the specific experiment they were used in. The hydrogels were then desiccated and stored until testing. Before each experiment, the hydrogels were rehydrated in saline overnight to impart ionic conductivity, and they were further cut to a specific shape if required by the study protocol.

The hydrogel samples, as well as their metal analogs, where appropriate, were affixed to the end of an irrigated 8-Fr ThermoCool SF ablation catheter (Biosense Webster, Irvine, CA) with a custom 3D-printed catheter attachment. This attachment allowed the hydrogel samples and their metal analogs to protrude beyond the attachment slightly (0.5–1 mm), allowing unimpeded contact with tissue. The attachment left the catheter’s irrigation ports unobstructed beside the hydrogel.

### Tissue Sample Staining and Imaging

To create 2% w/v staining solution, 2,3,5-triphenyltetrazolium chloride powder was dissolved in 0.9% saline solution and warmed to 37 °C. Ablated tissue samples were sliced vertically through the center of the ablation lesion and incubated for 20 minutes in the solution to enhance the margins between the ablated tissue and healthy tissue. Images of the stained ablated tissue were acquired after the samples were removed from the staining solution. Lesion depth and width were measured from the acquired images with ImageJ (v1.54f). Measurements were not blinded to electrode type.

### Characterizing Hydrogel Electrical Conductivity

Electrochemical impedance spectroscopy was used to measure the conductivity of the hydrogel (Figure S4). Hydrogel samples were centered between 2 electrodes and placed in a temperature controller set to physiological temperature. After physiological temperature was reached, an alternating current sweep was administered from 1 to 10^6^ Hz. The hydrogel resistance (*R*) was evaluated when the impedance reached a frequency of 500 kHz (RFCA frequency). Conductivity (σ) was determined by Equation 1, where *A* is the electrode area and *L* is the gel thickness.

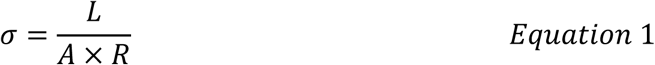

### Characterization of Lesion Geometry at Different Hydrogel Conductivities

Desiccated hydrogel plugs were swollen to equilibrium in either deionized water (Milli-Q; Millipore Sigma, Burlington, MA), resulting in plugs referred to as 0% saline plugs (00HP), or a sodium chloride aqueous (saline) solution. Selected concentrations for the experimental groups consisted of 0.5 wt% NaCl (05HP) and 1.5 wt% NaCl (15HP). Saline was used as a stand-in for altering the polymer chemistry of the hydrogel to change its electrical conductivity. Hydrogels were then cut to a diameter equal to that of the 8Fr ablation catheter tip, producing a 3.25 × 2.5-mm hydrogel plug (HP) electrode (Figure 1B).

Tissue samples were excised from the hearts by using a square cutting guide (3 × 3 cm) to partition consistent sections from the left ventricle, right ventricle, and septum. The tissue samples were then placed in a custom 3D-printed sample alignment cage before ablation. The tissue cages were placed in the saline tank atop the tissue stage, and the ablation catheter was slotted into the alignment cage guide.

Ablations (n=6 for each electrode type) were performed at a nominal power of 30 W for 30 seconds. Lesions formed by HP were compared to lesions created by a standard bare-metal (BM) ablation catheter (n=6).

### Characterization of Thermal Profile and Geometry of Lesions Formed by a Hydrogel Electrode

Left ventricular free wall and septal tissue samples were excised from the hearts. On each sample, ablation sites were selected such that the surface was free from trabeculations or steep contours. An initial incision was made with a scalpel on the surface of the tissue before inserting a custom 3D-printed thermistor array. The thermistors measured temperature at the surface of the tissue (T0) and at 2.5 mm (T1), 5.0 mm (T2), and 7.5 mm (T3) from the surface of the tissue. A custom interface was created to monitor and record temperatures from the thermistors.

Desiccated HPs were swollen to equilibrium in 0.9% saline to a final dimension of 3.25 × 2.5 mm; a metallic plug (MP) electrode of similar size (3.25 × 2.5 mm) was obtained. Ablations were performed with HP, MP, and BM at a contact force of 20 g at constant power set to either 20 W (n=12 for each electrode type) or 50 W (n=12 for each electrode type) (Figure 1C). Catheter irrigation with normal saline was maintained at a constant rate of 30 mL/min during ablation. Ablation was halted either when T2 crossed a temperature threshold of 55 °C or when 60 seconds had elapsed from the start of ablation. Data collection continued until T2 measured 40 °C after ablation was stopped. Peak temperatures were recorded from each of the thermistors, and the time to reach peak temperature from the start of ablation was calculated. Thermal latency was calculated as the difference in time between when the ablation was halted and when each thermistor reached peak temperature. At the end of each ablation, the tissue sample was stained and imaged as described previously. Additionally, overhead images of HPs were acquired using a DSLR camera under consistent lighting before and after each ablation to identify changes to the physical appearance of the hydrogel electrode.

### Characterization of Lesions Formed by Shaped Hydrogel Electrodes

To investigate how the shape of the hydrogel electrode affects lesion geometry, 2 differently shaped electrodes were created. Firstly, a hydrogel disk (HD) electrode was created by rehydrating molded desiccated hydrogel in 0.9% saline solution to a final cylindrical shape 10 mm in diameter and 2 mm in height (Figure 1E.1). Secondly, a hydrogel bar (HB) electrode was created by swelling a molded desiccated hydrogel in 0.9% saline solution to a final size of 30 × 5 × 2.5 mm (Figure 1E.2). A standard ablation catheter interfaced with the electrode at its center such that the face with the largest surface area faced the tissue surface. Ablations were performed for 30 seconds with a constant contact force of 20 g and constant irrigation of normal saline at 30 mL/min. Ablation power was set to 35 W with the disk electrodes and 30 W with bar electrodes. Lesions formed by the hydrogel electrodes were compared with those formed by a metallic equivalent (metal disk [MD] and metal bar [MB]) and by a standard ablation catheter (n=6 for each electrode type). For ablations with the bar electrodes, images of the lesion from the tissue surface were used to measure the horizontal and vertical dimensions of the lesions.

### Application of Hydrogel Electrodes to Form Atrial Lesions

Circular sections (20 mm) of atrial and esophageal samples were stacked such that the ablation electrode contacted the endocardial surface of the atrium and the epicardial atrial surface was in contact with the outer surface of the esophagus. The endoluminal side of the esophagus was in contact with a thermistor (Figure 1D) for continuous temperature monitoring.

Ablations (n=12) were performed with HP electrodes at a constant power of 30 W for 30 seconds. The catheter was irrigated with normal saline at a constant rate of 30 mL/min while a contact force of 20 g was maintained. Lesions created by HP ablation were compared with those created from a standard ablation catheter. Lesions were excised, imaged, and analyzed as described previously. Images of the HPs were taken before and after each ablation to identify any changes to the physical appearance of the hydrogel electrode.

### Steam Pop Frequency

Steam pop frequencies were recorded for all experiments within the experimental series. These frequencies were compared between the hydrogel and standard ablation catheter across all studies by a binary meta-analysis using a common effects model framework, using the Meta (v7.0-0) and Metasens (v1.5-2) packages in R (v4.4.0).

### Statistical Analysis

Continuous data are reported as mean ± SD. Differences in lesion geometry and temperature were analyzed by pairwise Wilcoxon rank-sum (Mann-Whitney U) tests with Holm correction for multiple comparisons in R (v4.4.0). P<0.05 was considered significant.

## Data Availability

The data that support the findings of this study are available from the corresponding author upon reasonable request.

## RESULTS

### Lesion Geometry at Different Hydrogel Conductivities

Twenty-four ablations were performed with the BM (n=6), 00HP (n=6), 05HP (n=6), and 15HP (n=6) (Figure 2A). Although no statistically significant differences in lesion geometry were found among the three hydrogel conductivities (Table 1), there was a trend of greater lesion size with higher saline concentration (Figure 3A, S2). The BM produced significantly deeper (*P*=0.030) and wider burns (*P*=0.041) than the 00HP and significantly wider burns than the 15HP (*P*=0.030).

**Figure 2.**
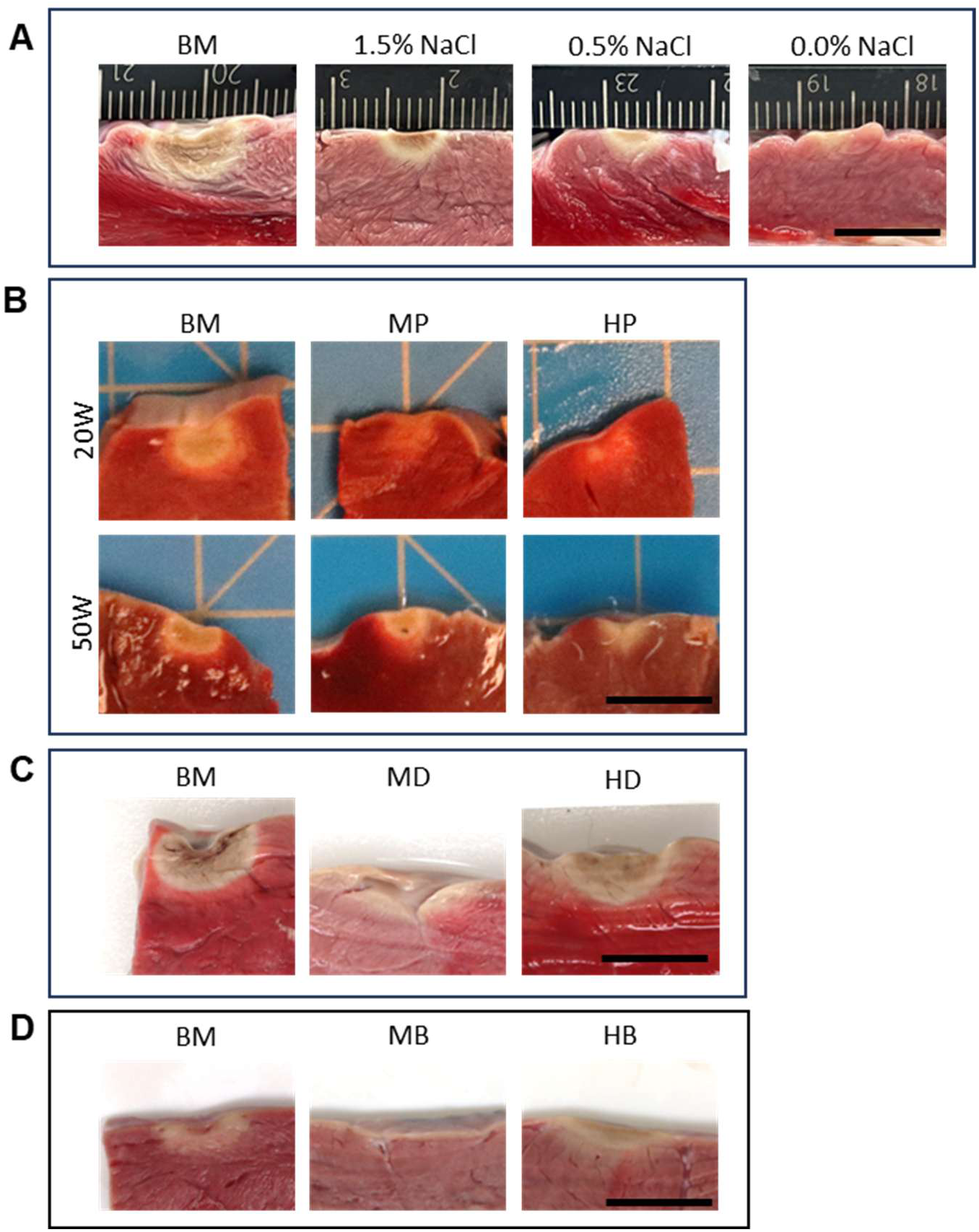
Representative images of lesion cross-sections from each test group. Lesions formed by each experimental electrode configuration were compared with those created by a standard catheter with a bare-metal (BM) electrode under the same ablation conditions. (**A**) Cross-sections of lesions formed by 2.5 × 3.25-mm round hydrogel electrodes at varying saline concentrations. (**B**) Cross-sections of lesions created by 2.5 × 3.25-mm round hydrogel plug (HP) and metal plug (MP) electrodes at 20 and 50 W power. Ablation was stopped when 60 seconds elapsed, or when T2 reached a temperature of 55°C, occurring in BM and MP at 50 W. (**C**) Cross-sections of lesions created by a 10 × 2-mm round hydrogel disk (HD) and round metal disk (MD) electrodes. (**D**) Cross-sections of lesions created by a 30 × 5 × 2.5-mm hydrogel bar (HB) electrode. Scale bars represent 10 mm. All images are scaled to the same size.

**Figure 3.**
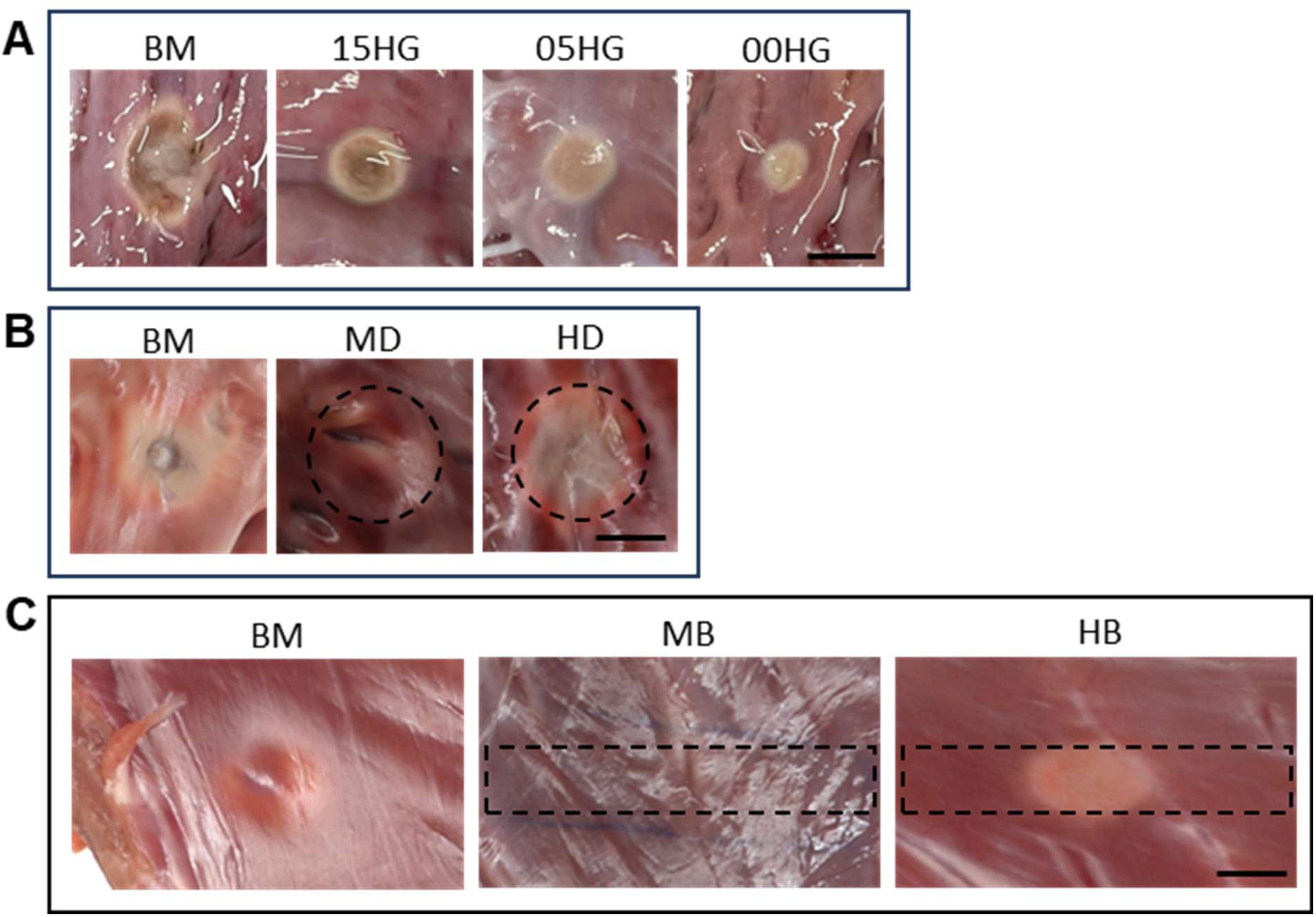
Representative images of surface lesions from each test group. Lesions formed by each experimental electrode configuration were compared with those created by a bare-metal (BM) standard ablation catheter under the same ablation conditions. (**A**) Surface lesion formed by 2.5 × 3.25-mm hydrogel plug (HP) electrodes at varying saline concentrations. (**B**) Surface lesions created by a 10 × 2-mm hydrogel disk (HD) electrode and a metal disk (MD) electrode of the same dimensions. (**C**) Surface lesions created by a 30 × 5 × 2.5-mm metal bar (MB) and hydrogel bar (HB) electrodes. Dashed lines denote where shaped electrodes contacted the tissue surface during ablation. Scale bars represent 5 mm. All images are scaled to the same size.

**Table 1.**
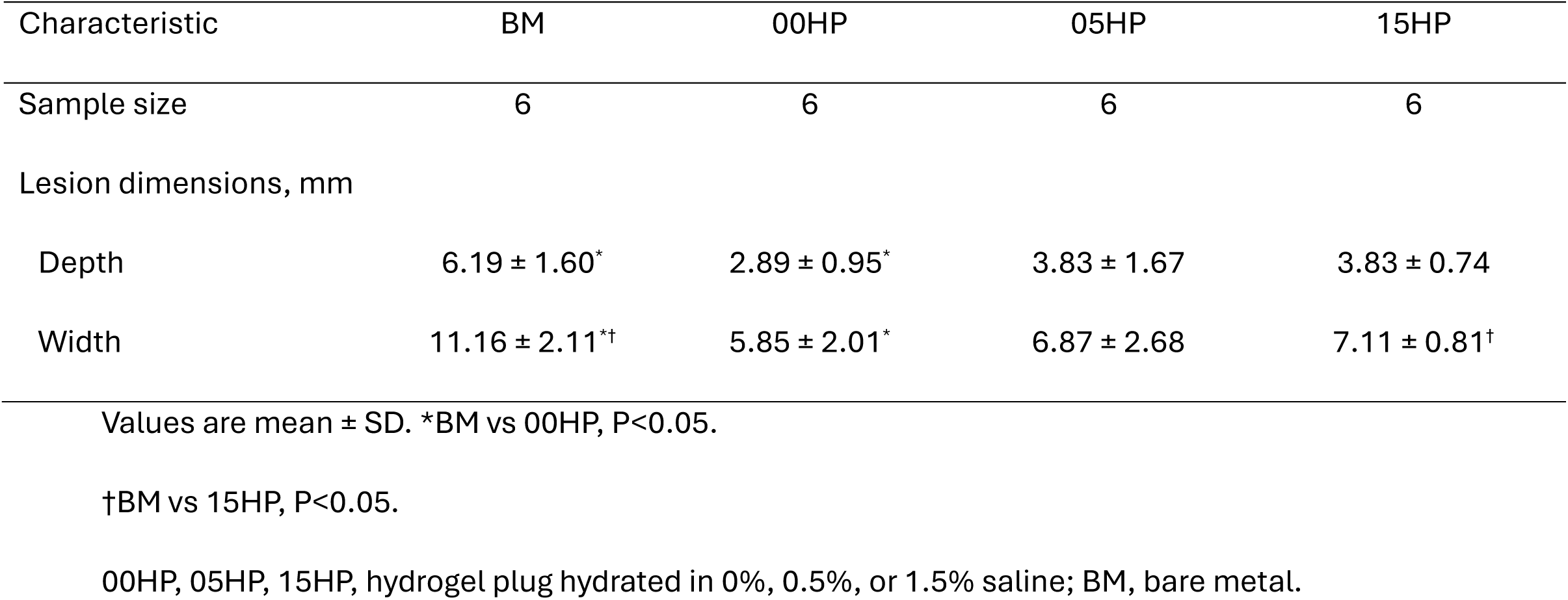
Characterization of Lesion Geometry With Varying Hydrogel Conductivity.

| Characteristic | BM | 00HP | 05HP | 15HP |
| --- | --- | --- | --- | --- |
| Sample size | 6 | 6 | 6 | 6 |
| Lesion dimensions, mm |  |  |  |  |
| Depth | 6.19 ± 1.60* | 2.89 ± 0.95* | 3.83 ± 1.67 | 3.83 ± 0.74 |
| Width | 11.16 ± 2.11*† | 5.85 ± 2.01* | 6.87 ± 2.68 | 7.11 ± 0.81† |

Values are mean ± SD. \*BM vs 00HP, P<0.05.
†BM vs 15HP, P<0.05.
00HP, 05HP, 15HP, hydrogel plug hydrated in 0%, 0.5%, or 1.5% saline; BM, bare metal.

### Thermal Profile of Lesions Created With Hydrogel Electrodes

In total, 72 ablations were performed with the BM, MP, and HP (n=24 for each) electrode types with ablation power set to either 20 or 50 W. Temperature data were excluded if the temperature probes failed during data collection: Data collected from T1 were excluded from 12/72 ablations, data collected from T2 were excluded from 2/72 ablations, and data collected from T3 were excluded from 5/72 ablations. Although care was taken to place T0 on the surface of the tissue, the thermistor was partly exposed to the saline tank in a few instances, which would have significantly cooled the thermistor. T0 was therefore excluded from all comparative analyses.

For the same electrode type, peak measured temperatures at T1 (T1_max_) and T2 (T2_max_) were significantly higher for ablation at 50 W than at 20 W (Table 2, Figure S3). Peak measured temperature at T3 (T3_max_) was not significantly different between ablation powers when HP or MP electrodes were used; there was a significant difference observed with a standard ablation catheter (peak temperature: 20W, 42.10 ± 1.66 °C; 50W, 44.65 ± 1.20 °C; *P*=0.039). There was no significant difference in T1_max_ and T2_max_ between electrodes at 20 W. However, when the ablation power was increased to 50 W, T1_max_ and T2_max_ were significantly lower for the HP electrode than for the MP or BM electrodes. Neither T1_max_ nor T2_max_ differed between BM and MP electrodes. T3_max_ was not significantly different across electrode groups at either of the ablation powers. Interestingly, peak temperatures seen with HP at 50 W did not differ from peak temperatures with MP or BM at 20 W (Table S3).

**Table 2.**
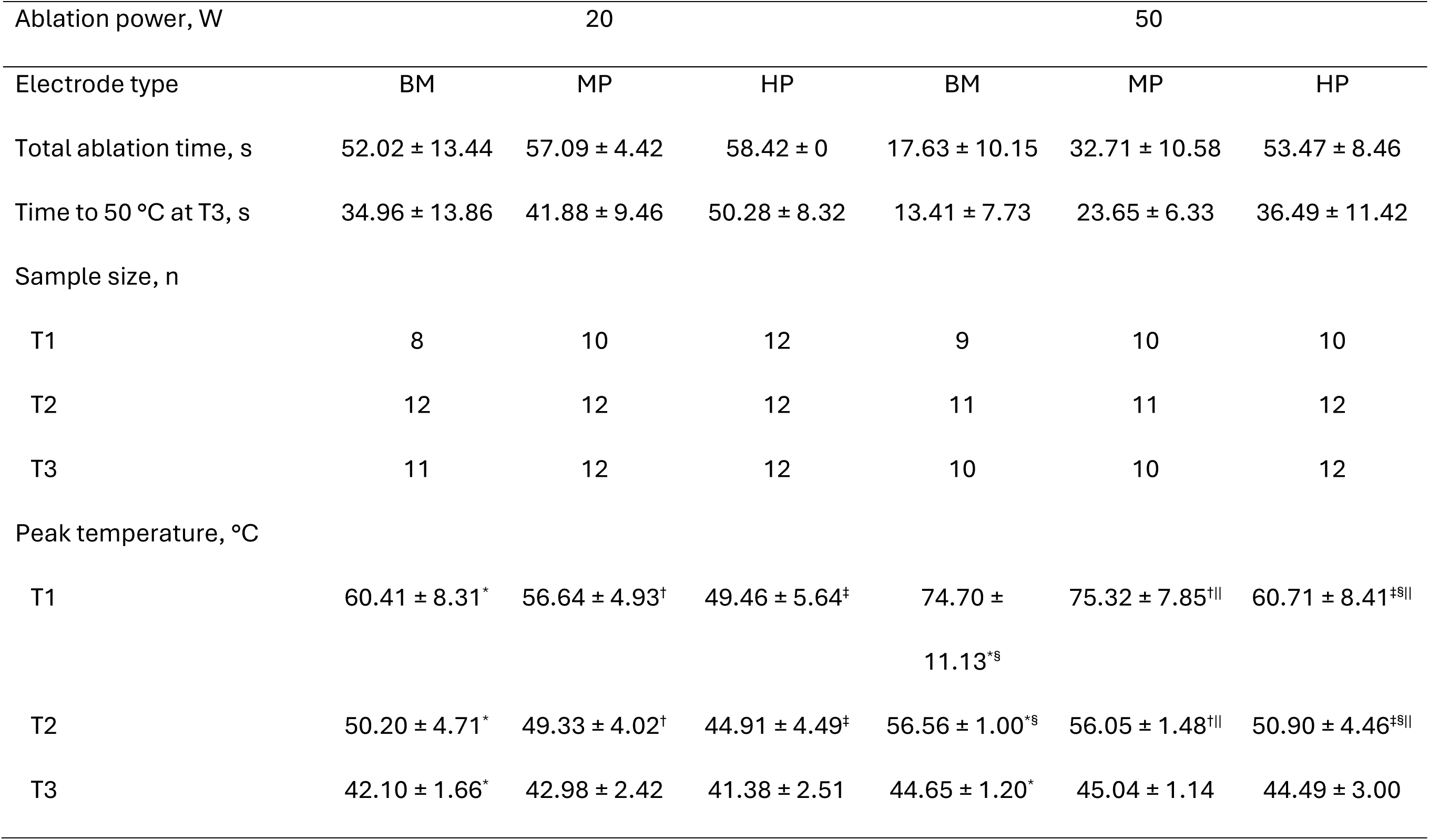
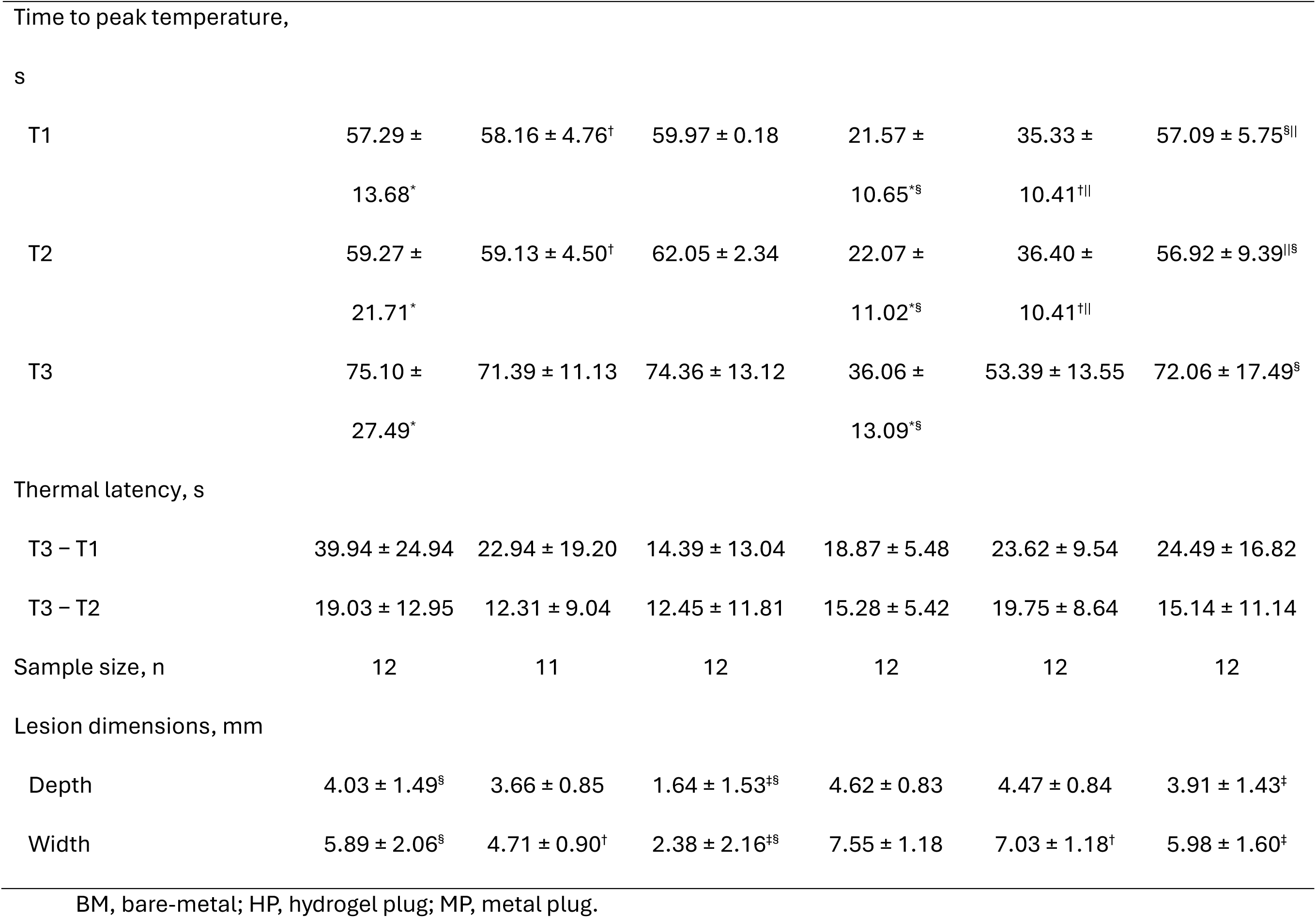
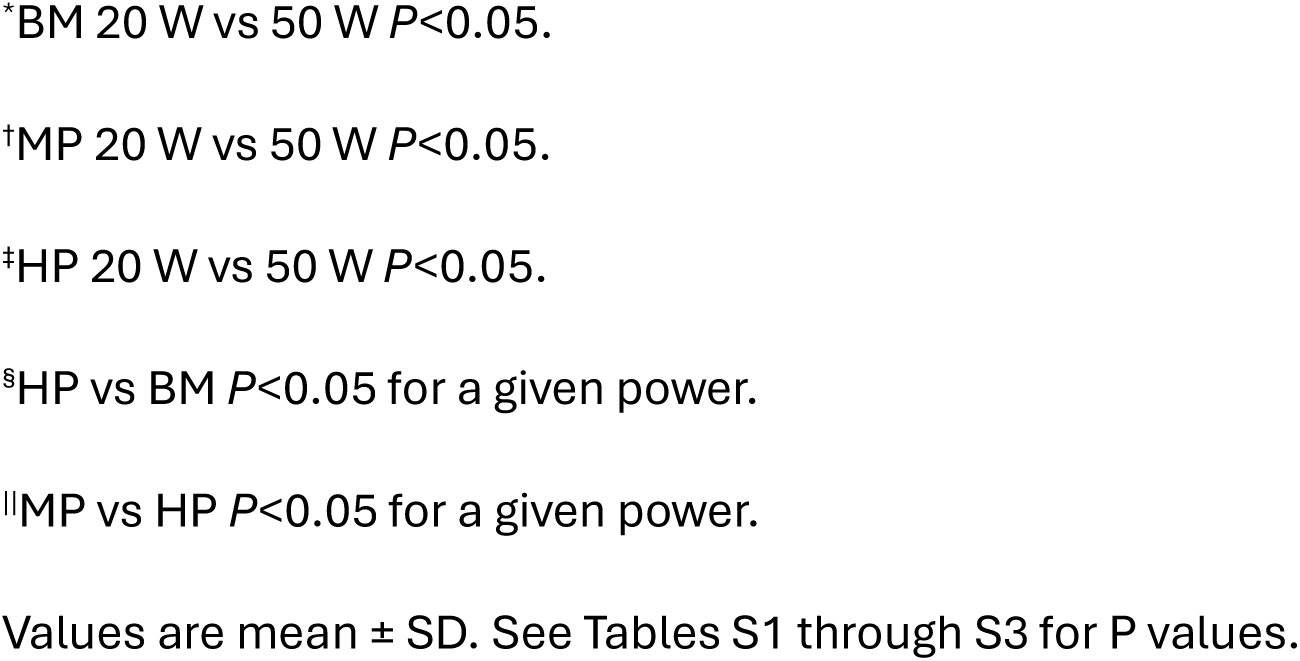
Thermal Profile and Lesion Geometry for Each Electrode Type.

At 20 W, only 3 ablations with BM and 1 ablation with MP reached the 55 °C threshold temperature (total ablation time: BM, 52.02 ± 13.44 s; MP, 57.09 ± 4.42 s; HP, 58.42 ± 0.00 s). At 50 W, all ablations with BM and MP and 4 ablations with HP reached the threshold temperature of 55 °C (total ablation time: BM, 17.63 ± 10.15 s; MP, 32.71 ± 10.58 s; HP, 53.47 ± 8.46 s).

Regarding time to peak temperature from start of ablation, ablations at 20 and 50 W did not differ in time to peak temperature at T1, T2, or T3 when the HP electrode was used. During ablations at 50 W with MP and BM, T1 and T2 reached 50 °C significantly faster than when using HP (*P*=0.0046 and *P*=0.0058 for BM and MP at T1 and *P*=0.0013 and *P*=0.0057 for BM and MP at T2, respectively). The BM electrode reached 50 °C fastest of the 3 electrode types at the T2 thermistor. This was the case regardless of the ablation power used.

When lesion geometry was compared between HP and BM ablations at 20 W (Figure 2B), HP lesions were shallower (1.64 ± 1.53 mm vs 4.03 ± 1.49 mm, *P*=0.044) and narrower (2.38 ± 2.16 mm vs 5.89 ± 2.06 mm, *P*=0.026). At 50 W, lesions formed by BM, MP, and HP electrodes did not differ significantly in geometry (Table 2).

### Shaped Hydrogel Disk Electrode

A total of 18 ablations were performed with the BM, HD, and MD (n=6 for each). Lesions formed by the HD (Figure 2C, 3B) were not significantly different from those formed by BM in width (9.47 ± 0.89 mm vs 11.41 ± 2.39 mm, *P*=0.383) or depth (4.54 ± 0.90 mm vs 4.84 ± 0.55 mm, *P*=0.575). Additionally, lesions formed with the MD were shallower than those produced by the BM (2.30 ± 1.70 mm vs 4.84 ± 0.55 mm, *P*=0.015), whereas lesion widths were not significantly different (5.84 ± 4.57 mm vs 11.41 ± 2.39 mm, *P*=0.383) (Table 3).

**Table 3.** Characterization of Lesion Geometry With Differently Shaped Electrodes.

| Characteristic |  | BM | MD | HD | BM vs MD (*P*) | BM vs HD (*P*) | MD vs HD (*P*) |
| --- | --- | --- | --- | --- | --- | --- | --- |
| Disk electrode | Sample size, n | 6 | 6 | 6 |  |  |  |
|  | Lesion dimensions, mm |  |  |  |  |  |  |
|  | Depth | 4.84 ± 0.55 | 2.30 ± 1.70 | 4.54 ± 0.90 | 0.015 | 0.575 | 0.061 |
|  | Width | 11.41 ± 2.39 | 5.84 ± 4.57 | 9.47 ± 0.89 | 0.383 | 0.383 | 0.470 |
|  | Characteristic | BM | MB | HB |  | BM vs HB (*P*) |  |
| Bar electrode | Sample size, n | 6 | — | 6 |  |  |  |
|  | Lesion dimensions, mm |  |  |  |  |  |  |
|  | Depth | 4.82 ± 1.11 |  | 3.73 ± 0.49 |  | 0.063 |  |
|  | Width | 8.71 ± 1.65 |  | 10.51 ± 1.40 |  | 0.031 |  |

Values are mean ± SD. MB electrodes produced no lesions.
BM, bare metal; HB, hydrogel bar; HD, hydrogel disk; MB, metal bar; MD, metal disk.

### Shaped Hydrogel Bar Electrode

A total of 18 ablations were performed with the BM, MB, and HB (n=6 for each). The MB electrodes produced no lesions (Figure 3C). In contrast, the HB electrodes (Figure 2D) produced lesions with greater maximum widths than those produced by the BM (10.51 ± 1.40 mm vs 8.71 ± 1.65 mm, *P*=0.031). However, no significant differences were found in lesion depth (3.73 ± 0.49 mm vs 4.82 ± 1.11 mm, *P*=0.063) (Table 3). The lesions produced by the HB were ellipsoidal, as shown by the difference between their long and short axes (11.15 ± 0.60 mm vs 6.52 ± 0.69 mm, *P*=0.031), with an aspect ratio of 1.72 ± 0.19. In contrast, lesions formed by the BM were circular.

### Atrio-Esophageal Tissue Stack Study

The HP atrial lesions were significantly narrower than the BM lesions (8.68 ± 2.34 mm vs 12.56 ± 2.88 mm, *P=0.002*; Table 4). Lesion widths measured from esophageal tissue also revealed that HP electrodes produced smaller lesions than BM electrodes (3.05 ± 2.35 mm vs 8.70 ± 2.69 mm, *P*<0.001) (Figure 4). All ablations created transmural lesions across the atrium. Total lesion depth, which was calculated as the sum of lesion depths across the atrial and esophageal walls, was shallower with HP than with BM (4.03 ± 1.25 mm vs 7.58 ± 0.78 mm, *P*<0.001). The esophageal lumen temperature was lower during HP ablation than during BM ablation (56.33 ± 3.80 °C vs 61.30 ± 5.14 °C; *P=0.04C* ).

**Figure 4.**
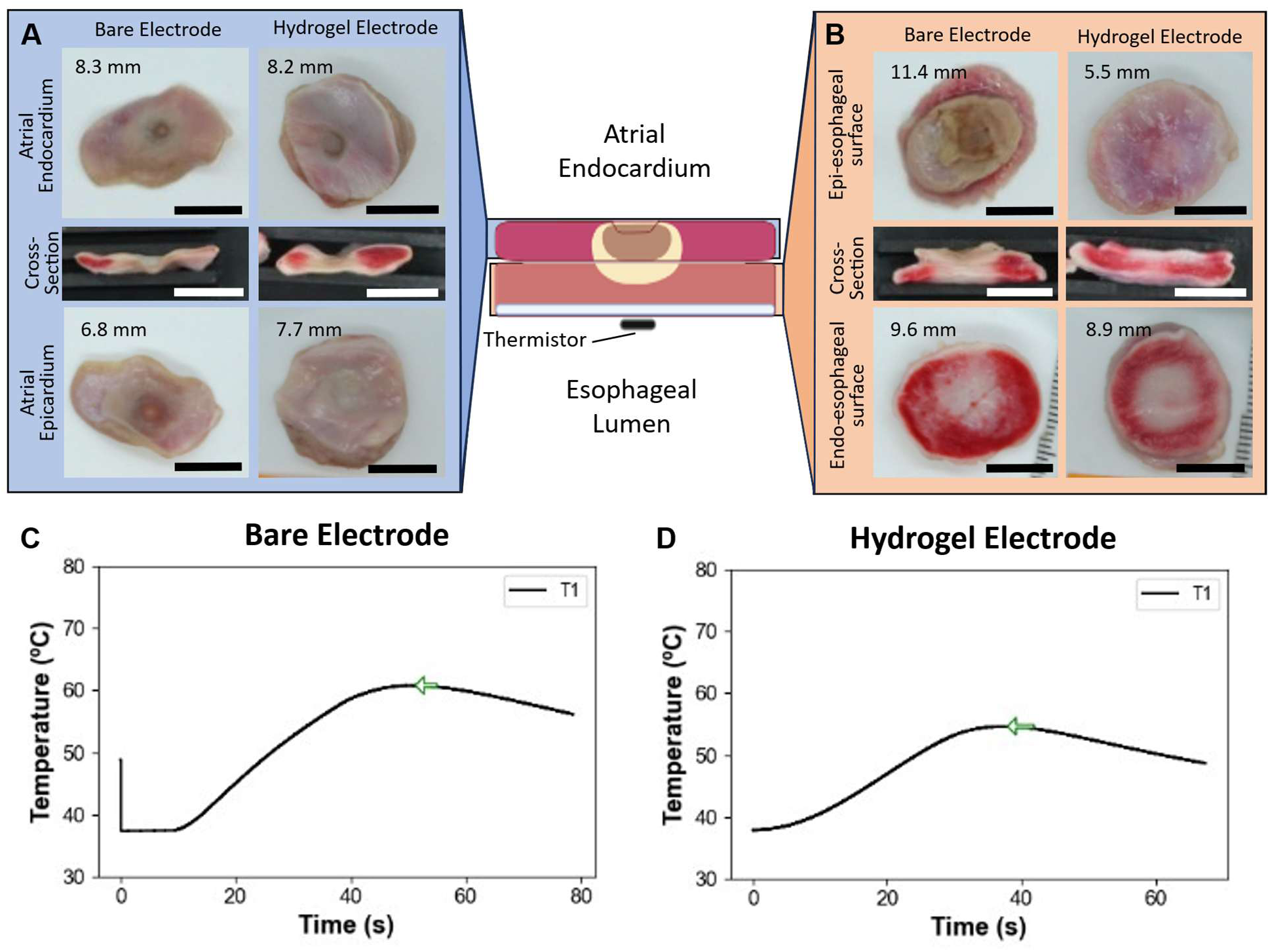
Representative atrioesophageal stack ablation gross histology samples and temperature profiles from each group. (**A**) Representative atrial tissue samples after ablation using the BM electrode and 2.5 × 2.0-mm round hydrogel plug electrodes, and (**B**) corresponding representative esophageal tissue samples; lesion widths are noted for each sample. (**C**) Representative temperature profile recorded from the endoluminal thermistor during ablation with the BM electrode. (**D**) Representative temperature profile recorded during ablation with the hydrogel plug electrode. Peak temperature is marked with the green arrow. Scale bars represent 10 mm. All images are scaled to the same size.

**Table 4.** Atrioesophageal Stack Study Lesion and Temperature Measurements.

| Characteristic | BM | HP | *P* Value |
| --- | --- | --- | --- |
| Lesion dimensions, mm |  |  |  |
| Atrial lesion width | 12.56 ± 2.88 | 8.68 ± 2.34 | 0.002 |
| Esophageal lesion width | 8.70 ± 2.69 | 3.05 ± 2.35 | <0.001 |
| Total lesion depth | 7.58 ± 0.78 | 4.03 ± 1.25 | <0.001 |
| Esophageal lumen temperature |  |  |  |
| Peak temperature, °C | 61.30 ± 5.14 | 56.33 ± 3.80 | 0.046 |
| Time to peak temperature, s | 40.13 ± 9.18 | 37.31 ± 6.83 | 0.560 |

Values are mean ± SD. BM, bare metal; HP, hydrogel plug.

### Steam Pops

In all tests of different hydrogel conductivities, only 2 steam pops occurred; both happened when BM was used. No steam pops occurred during the hydrogel thermal profile tests. In the atrio-esophageal tissue stack study, 2 steam pops occurred during ablation with the BM electrode, whereas no steam pops occurred with the HP electrode. In the 1-cm HD study, 2 steam pops occurred with the HD electrode, and 4 occurred with the BM electrode. In the HB study, the BM and HB electrodes produced 1 steam pop each. No steam pops were recorded for any of the metal analogs (MP, MD, MB). The meta-analysis found a lower relative risk ratio (0.34; 95% CI 0.13–0.88) for steam pop formation with hydrogel electrodes versus BM electrodes (Figure S5).

### Hydrogel Characterization After Ablation

Small air pockets were observed in some of the hydrogel electrodes after ablation (Figure S6): 2 (of 6) hydrogel electrodes in the 00HP group, 5 (of 6) in the 05HP group, and 4 (of 6) in the 15HP group. Eight of 12 hydrogel electrodes formed visible bubbles after ablation at 50 W, while no bubbles were observed after ablation at 20 W. One HD had an indentation where the catheter contacted the hydrogel electrode, but no air bubbles were observed. Finally, no changes were observed in the HB electrodes after ablation.

## DISCUSSION

In this *ex vivo* study, a conductive hydrogel served as a moldable RFCA electrode medium that produced lesion geometries its rigid metallic analogs could not. The clearest demonstration was the bar electrode: a hydrogel bar generated wide, ellipsoidal lesions, whereas a size-matched metal bar produced no lesion at all. The disk experiments make the same point in a complementary way—a hydrogel disk produced lesions that did not differ significantly from those of a standard bare-metal electrode in depth or width, while an equivalently sized metal disk fell significantly short. Taken together, these findings indicate that it is the hydrogel medium, rather than electrode dimensions alone, that enables large, contiguous, geometrically defined lesions. Because the gel can be molded to a target footprint and conforms to the tissue surface, it offers a means of tailoring lesion shape that is not available with fixed metallic electrodes.

The plug experiments characterize the biophysics underlying this behavior rather than serve as a performance comparison the hydrogel is intended to win. Across the conductivity series, lesion size trended with ionic content, confirming that ions are the principal—though not sole—charge carrier and that conductivity is tunable through hydration. The thermal-profiling data show that the hydrogel attenuates energy delivery: subsurface peak temperatures were lower and total ablation times longer with the hydrogel plug than with bare or metallic plugs, and peak temperatures with the hydrogel at 50 W approximated those of bare metal at 20 W. This attenuation is precisely why the plug produced shallower, narrower lesions than bare metal—an expected consequence of the medium’s far lower conductivity, and a property to be exploited rather than a deficiency to be corrected.

Framed by depth regime, energy attenuation is a liability and an asset depending on the target. For deep, intramural ventricular substrate, reduced energy transfer would limit reach, and a hydrogel electrode in its present formulation would not be the appropriate tool. For atrial ablations, where transmurality must be achieved without injuring adjacent structures, the same property provides a protective margin. The atrioesophageal model captured this directly: hydrogel ablation achieved fully transmural atrial lesions while simultaneously reducing esophageal lumen temperature, total lesion depth, and esophageal lesion width relative to bare metal. The clinically meaningful endpoint— transmural atrial injury—was reached, while the off-target thermal footprint was reduced. The lower steam-pop risk observed with hydrogel electrodes across experiments is consistent with this gentler thermal behavior and reinforces the safety signal. These results reframe the value of a conductive hydrogel electrode away from raw lesion depth and toward two capabilities that rigid electrodes lack: control over lesion geometry and a more favorable collateral-injury profile. The biophysical basis for both warrants closer examination.

### Biophysics and Limitations of Radiofrequency Ablation

Ablative RF energy heats up tissue during an initial, resistive heating phase and then a conductive heating phase as thermal energy dissipates from the surface into the tissue.^11^ This effect can produce inconsistent lesions, which frequently necessitates repeat burns; also, incomplete lesions can introduce new reentrant circuits.^12^ This becomes especially problematic during procedures that necessitate the creation of long, contiguous lesions, such as pulmonary vein isolation procedures, which require multiple closely spaced ablations.

### Effect of Hydrogel Conductivity on Lesion Formation

We observed that increasing the saline concentration in the hydrogel electrodes—from deionized water (00HP) to 0.5% saline (05HP) and 1.5% saline (15HP)—resulted in a nonsignificant trend toward larger lesions. This trend suggests that higher ionic content enhances the hydrogel’s electrical conductivity, facilitating greater energy transfer to the tissue. The ions in the hydrogel are the primary carrier of energy through the gel but not the sole one, as evidenced by the formation of lesions even with the 00HP electrodes. However, the hydrogel has substantially lower conductivity than metals like platinum (∼2 S/m vs ∼9.43 × 10^6^ S/m),^13^ which attenuates energy delivery. Reducing conductivity does not, by itself, improve lesion formation: a standard catheter with appropriate parameters can match the lesion depth of a similarly sized hydrogel electrode on flat tissue. The value of the hydrogel lies elsewhere—in its capacity to be molded into lesion-shaping geometries that a rigid electrode cannot form, and in the accompanying energy profile that we exploit for esophageal sparing.

### Thermal Profiles and Accounting for Energy Attenuation

To further study the biophysics of ablation with a hydrogel electrode, we used a probe to measure temperature within the tissue as it was ablated. As part of this investigation, we chose to ablate until 1 of 2 conditions were met: Either the temperature recorded 5 mm below the surface reached at least 55 °C (cardiac cells die when heated beyond 50 °C^14^), or at least 60 seconds had elapsed from the start of the ablation. Adhering to these limits enabled us to study the thermal transfer through different electrode types and to compare lesion sizes after a target temperature was achieved. In all electrode types, when ablative energy was delivered to the tissue, the temperature of the tissue nearest to the electrode (at T1 and T2) increased rapidly, suggesting that resistive heating was primarily responsible. In contrast, the rate of temperature change more distal to the electrode (at T3) was comparatively slow, suggesting that conductive heating played a larger part.

Evidence suggests that a longer electrode sinks more energy into the surrounding environment and thus produces smaller lesions^15^. To control for the greater total electrode length resulting from appending a hydrogel electrode to the end of the catheter electrode, we used matching metal electrodes as an additional control. Unlike Nakagawa et al,^15^ we did not observe any significant differences in lesion profiles with the use of a longer metallic electrode (MP) than with a shorter one (BM). This is probably due to the stop conditions (ie, 55 °C or 60 s) we used in each ablation. Although T1_max_ and T2_max_ did not differ significantly between the MP and BM, the BM reached 55 °C at T2 about 15 seconds faster than MP. The difference in total ablation time can be attributed to a longer conductive electrode sinking some ablative energy to the environment. Compared with BM and MP, HP had a lower T1_max_ and T2_max_ and a longer total ablation time, indicating that the hydrogel attenuates the ablative power even more than the metal. While a portion of the ablative energy is probably sinking into the ambient saline environment along the length of the hydrogel electrode, a larger portion of it is probably attenuated by the greater impedance the hydrogel electrode imparts.

### Air Pocket Formation Within the Hydrogel Electrodes

Structural changes to the hydrogel, like the formation of air pockets within the hydrogel during ablation, suggest that the hydrogel itself is undergoing resistive heating, evidenced by the greater incidence of air-pocket formation in 50-W ablations than in 20-W ablations. This resistive heating could be enough to vaporize water existing within the hydrogel matrix, resulting in the air pockets seen. While precise temperature profiles within the hydrogel electrodes remain unclear, bubble formation could affect energy delivery and electrode integrity. However, despite air-pocket formation and several tissue steam pops, all of the hydrogel electrodes remained intact. Additionally, the occurrence of air-pocket formation within the hydrogel could be related to the lower relative risk ratio of steam pop during ablation with the hydrogel electrodes. Further studies should investigate this phenomenon to optimize hydrogel formulations and ablation parameters, ensuring consistent performance and safety.

### Shaping Lesion Geometry With Hydrogel Electrodes

Conductivity reduction is not itself the therapeutic mechanism; the benefit comes from shapeability and the energy profile it produces. Despite its lower conductivity, the hydrogel created fully transmural lesions in atrial applications. Additionally, these electrodes can be easily molded into different shapes to form lesions of various dimensions. Although catheters like the Sphere-9 (Medtronic, MN) can create larger lesions through their nitinol basket electrode geometry^5,16^, their metallic electrodes are too rigid to conform well to the anatomy of the heart and, especially, to reach recessed locations. In contrast, the mechanical compliance of the hydrogel enables it to conform readily to the tissue (Figure S7). This compliance is also advantageous in designing a more flexible, steerable catheter. In the present study, hydrogel electrodes produced lesions of different shapes; for example, the HB produced ellipsoidal lesions with an aspect ratio of 1.72; these lesions were wider along the long axis and shallower than those formed by the standard (BM) ablation catheter. The HB formed lesions whose short axis was limited to within 1.3 mm of the hydrogel electrode’s edge. The HD created lesions closely matching the disk’s dimensions. These findings suggest that hydrogel electrodes can direct RF energy more precisely than a standard catheter, allowing clinicians to tailor lesion shapes to specific anatomical and clinical needs. This capability is particularly advantageous when one is ablating near sensitive structures or in regions with complex geometries, such as the atrial appendage or the tissue around the papillary muscles.

### Clinical Implications and Advantages

Beyond shaping, the hydrogel’s capacity to form contiguous lesions in a single application could, if borne out *in vivo*, reduce the number of sequential burns needed for linear lesion sets and the associated procedure time. More importantly, the hydrogel improved the ratio of on-target to off-target heating rather than merely lowering delivered energy: in the atrioesophageal model it produced fully transmural atrial lesions while simultaneously yielding significantly lower esophageal lumen temperatures than BM electrodes. Because reducing a bare electrode’s power would lower esophageal temperature only at the cost of atrial transmurality, this simultaneous result reflects a widened therapeutic window—an advantage particularly relevant to ablation near sensitive structures such as the esophagus.

### Limitations

The ex vivo model, while controlled, does not replicate the dynamic environment of a living heart, where factors like blood flow, tissue perfusion, and physiological movement can affect ablation outcomes. Additionally, the electrode-hydrogel attachments were not fully optimized in terms of interface uniformity, potentially influencing energy transfer efficiency and mechanical stability. Variations in tissue properties and electrode-tissue contact may have contributed to the observed variability in lesion sizes. The scope of the study did not extend to creating a method to deliver hydrogel electrodes of various sizes to a target location.

### Future Directions

Further research is needed to refine hydrogel electrode technology for clinical application. In future studies, the hydrogel will be directly bonded to the electrode, and we will test different hydrogel electrode geometries to optimize the shaping and energy delivery of the system. Additionally, further development of the hydrogel’s chemistry will enable the creation of even more compliant hydrogel electrodes that can be deployed and retracted through a standard-size delivery sheath, making intravenous ablation with these larger hydrogel electrodes viable. In vivo pre-clinical studies will be essential to validate the effectiveness and safety of hydrogel electrodes in the clinical setting.

### Conclusions

This work shows that conductive hydrogel electrodes can be used to produce lesions when used in RFCA. The hydrogel electrodes reduce the overall energy delivered to the tissue, with a high degree of modulation based on their electrical conductivity. Importantly, the flexible and highly shapeable gel can be used to control the geometry of the lesion and better define its boundaries. This feature makes hydrogel electrodes useful for applications requiring a larger contact area, such as making linear burns; for producing large, contiguous lesions with more uniform properties; and for use in spaces with difficult tissue convolutions where the hydrogel could conform better to the tissue geometry.

## Data Availability

The data supporting the findings of this study are available from the corresponding author upon reasonable request.

## Acknowledgments

The authors thank Stephen N. Palmer, PhD, ELS, of the Department of Scientific Publications at The Texas Heart Institute at Baylor College of Medicine for his editorial contributions and Kavya Sinha, MD, of the Electrophysiology Clinical Research and Innovations group at The Texas Heart Institute at Baylor College of Medicine for her review of the manuscript.

## Sources of Funding

This work was supported by the contributions from the Cizik Foundation.

## Disclosures

Drs Cosgriff-Hernandez, Post, and Razavi and Mathews John are inventors on a patent application covering electrically conductive hydrogel electrodes (US12427284B2). Drs Cosgriff-Hernandez, Post, and Razavi are founders of Rhythio Medical Technologies, which seeks to commercialize hydrogel electrodes, and Mathews John provides consulting services to Rhythio Medical Technologies. The other authors report no conflicts.

## Nonstandard Abbreviations and Acronyms

00HP, 05HP, 15HP: hydrogel plug hydrated in 0%, 0.5%, or 1.5% saline
BM: bare-metal
HB: hydrogel bar
HD: hydrogel disk
HP: hydrogel plug
MB: metal bar
MD: metal disk
MP: metal plug
RFCA: radiofrequency catheter ablation

## Supplemental Material

**Figure S1.**
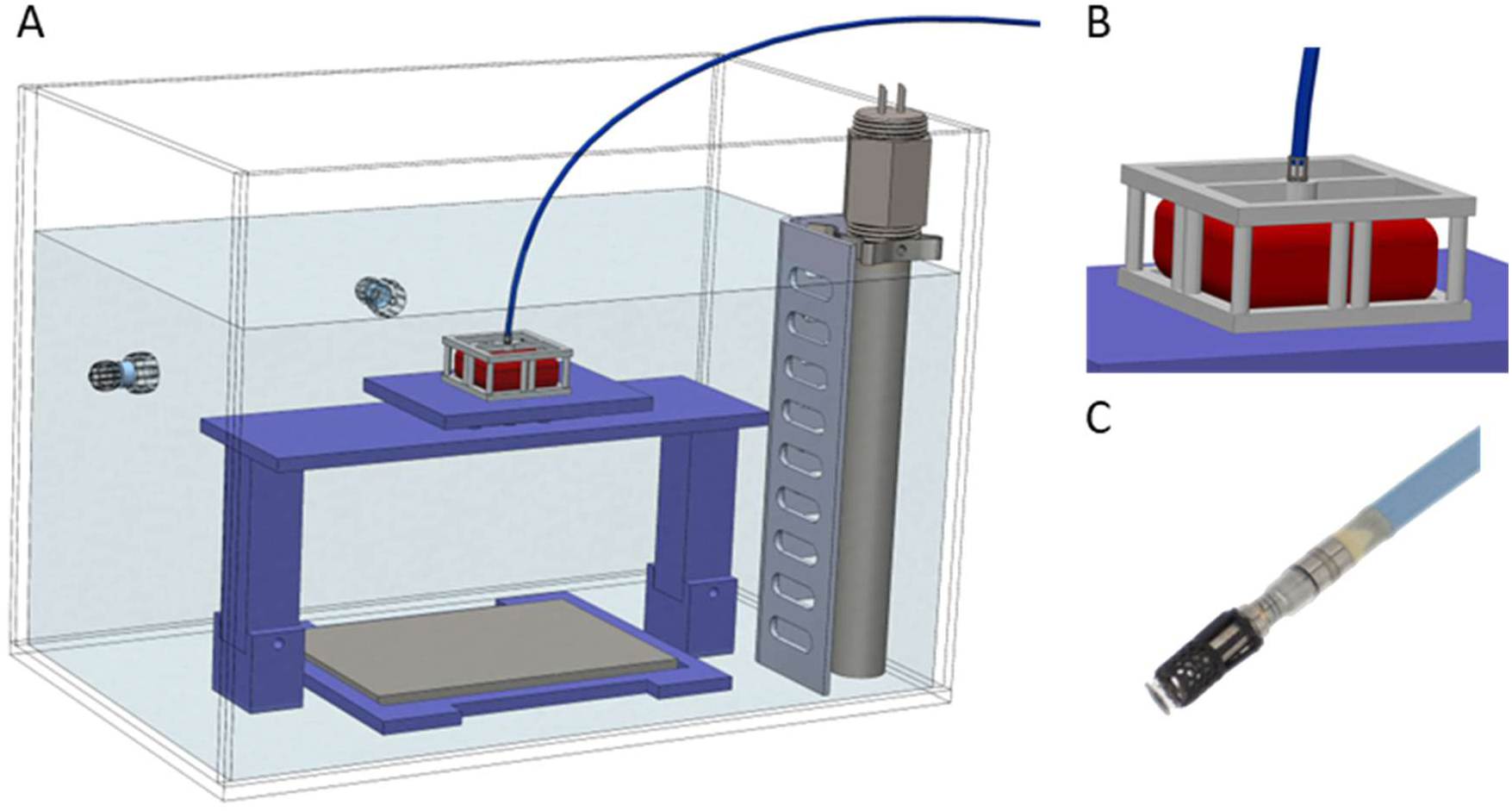
(**A**) Full tank assembly for ex-vivo tissue ablation, including tissue stage, grounding pad, circulating water heater set to 37C, and 0.9% saline solution. (**B**) Custom designed and printed tissue holder for ablation catheter alignment. (**C**) Photo of custom designed and printed hydrogel punch holder fitted to the end of an RF ablation catheter.

**Figure S2.**
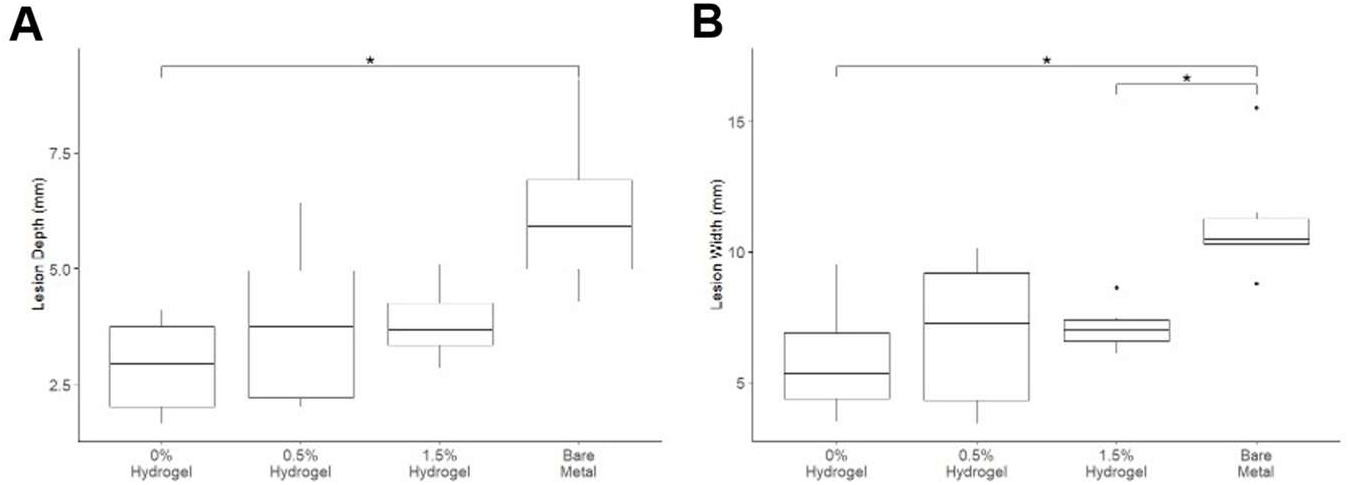
(**A**) Measured lesion depth from cross-sectional images. (**B**) Measured lesion width from cross-sectional images.

**Figure S3.**
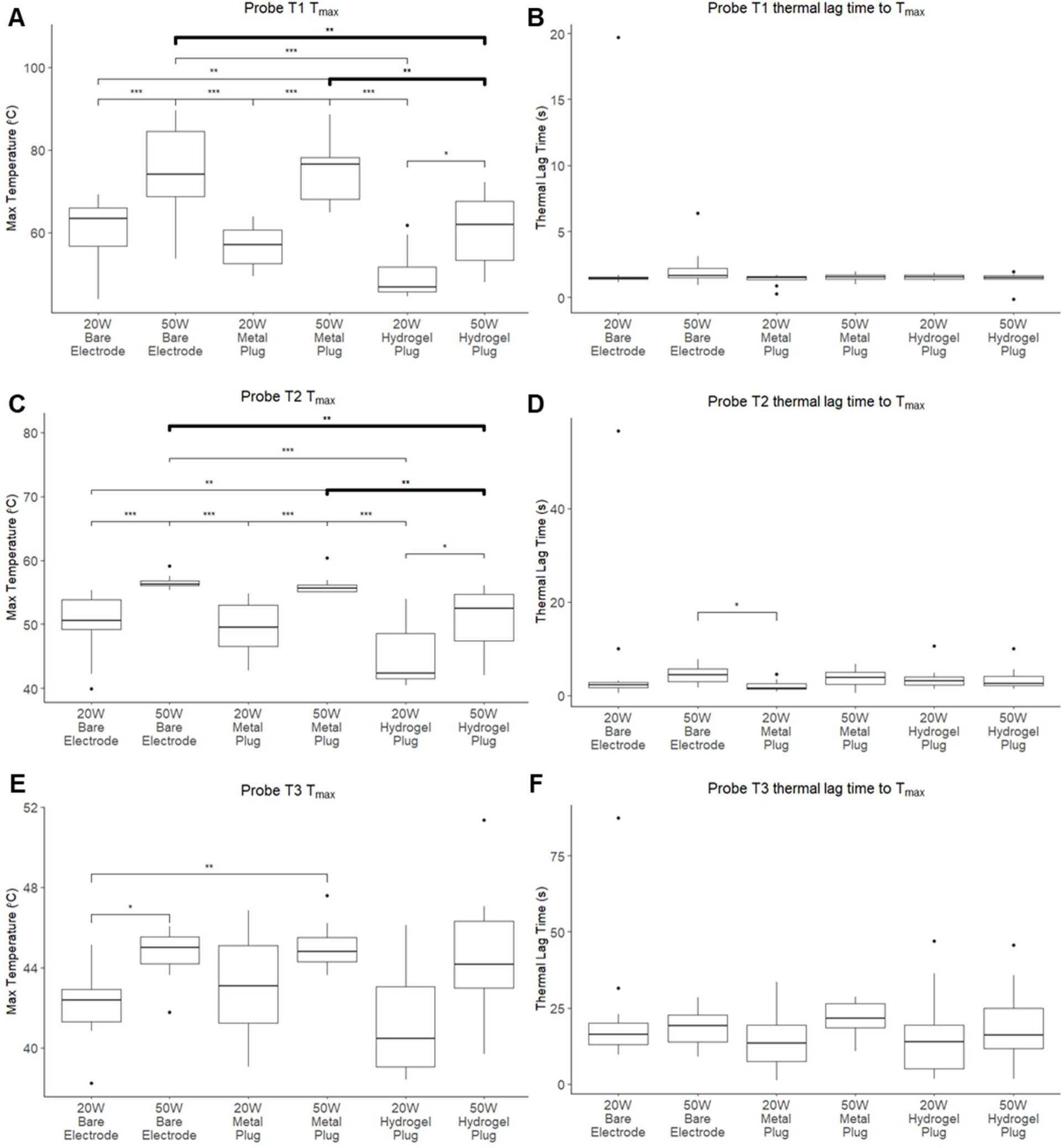
Average of peak temperature measurements from (**A**) T1 at 2.5mm depth, (**C**) T2 at 5mm depth, and (**E**) T3 at 7.5mm depth below the surface of the tissue. Time to peak temperature calculated from the end of ablation at (**B**) T1, (**D**) T2, and (**F**) T3.

**Figure S4.**
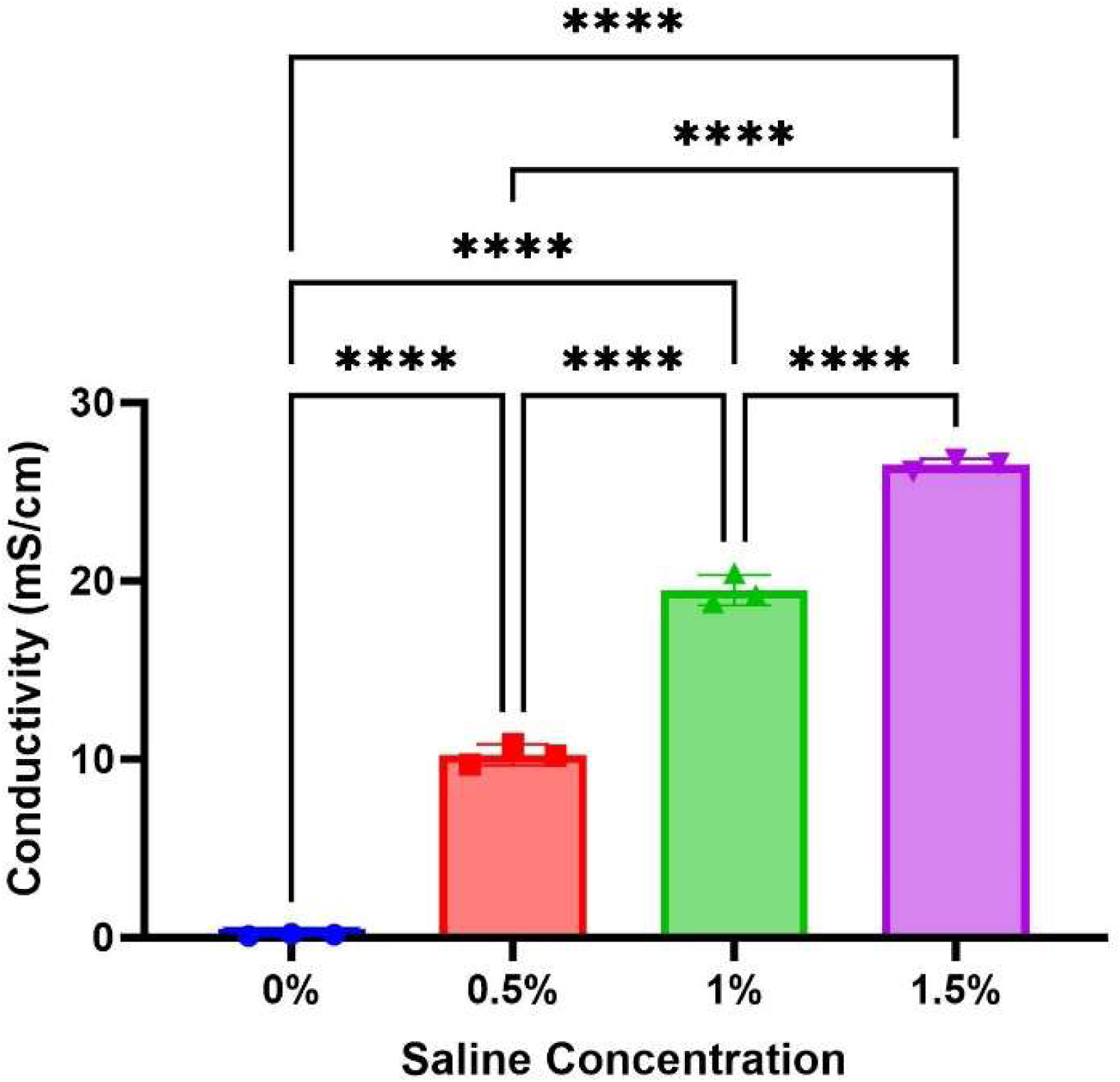
Conductivity of hydrogels at physiological temperature as a function of salt content. Hydrogel conductivity at 500 kHz was assessed using electrochemical impedance spectroscopy. Individual values, mean and standard deviation are displayed (ns = not significant; * = p < 0.05; **** = p < 0.0001).

**Figure S5.**
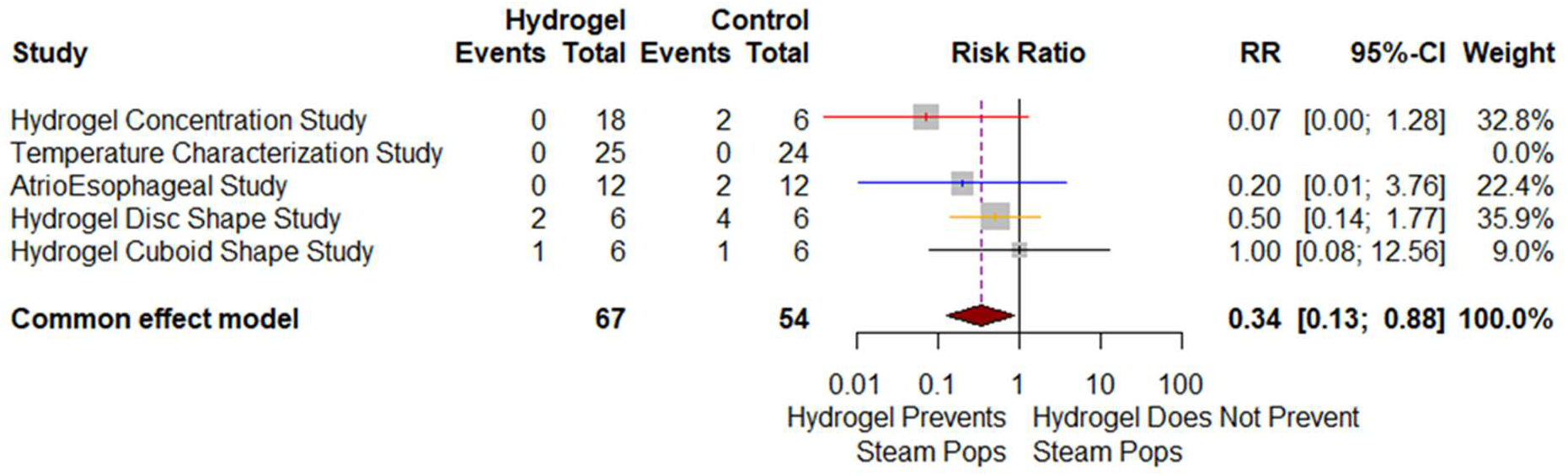
Forest plot of relative risk ratio for steam pop formation between hydrogel and bare electrode conditions.

**Figure S6.**
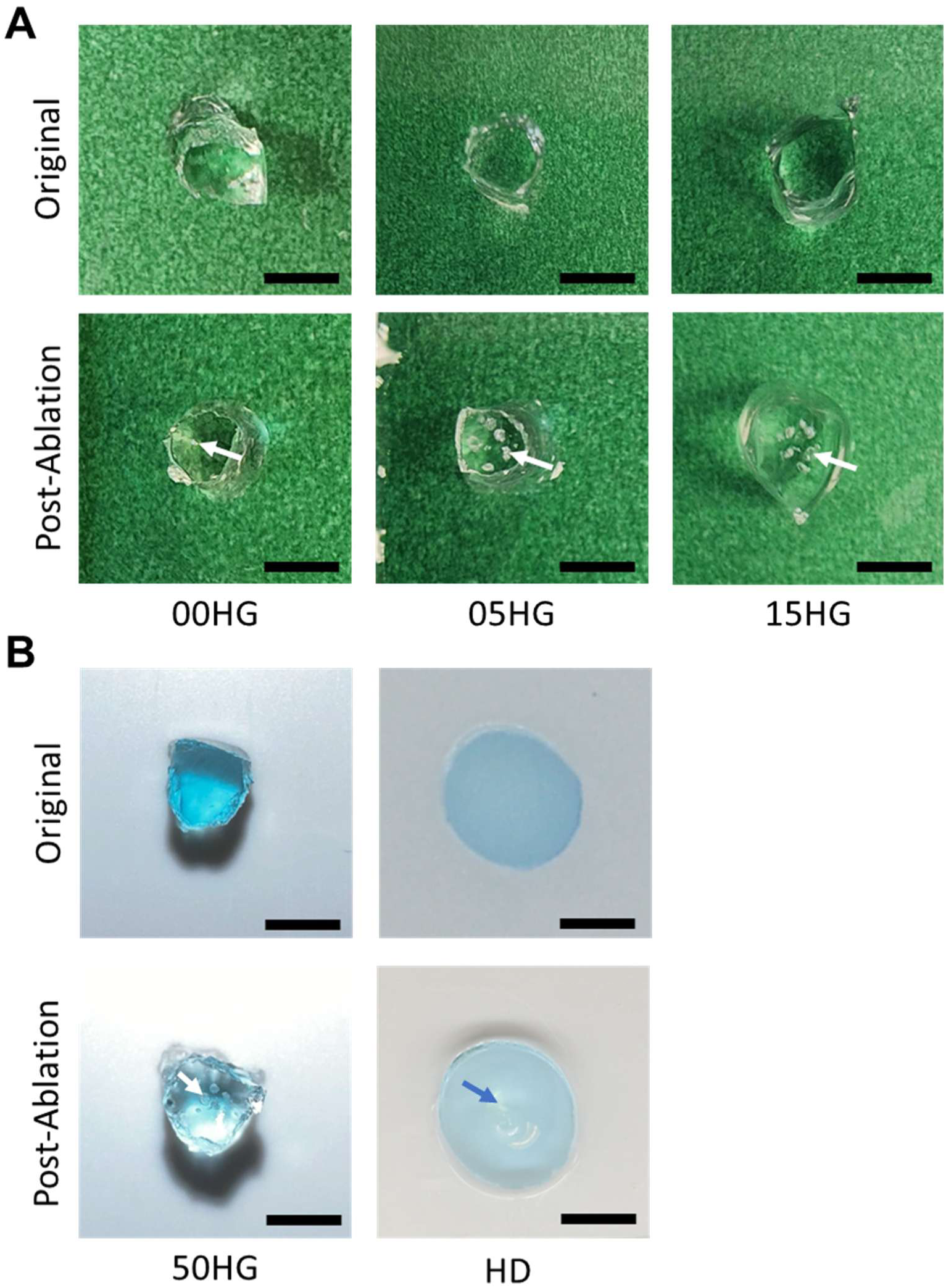
Changes in the physical appearance of several hydrogel electrode samples. (**A**) Formation of air bubbles (white arrows) in hydrogel samples before after ablation while characterizing hydrogels with varying conductivity. Scale bars represent 2mm. (**B**) Noted visual changes during further characterization, including the formation of air bubbles (white arrow) at 50W and the formation of an indentation in the hydrogel disk from the catheter at 20W. HP (50 W) scale bar represents 2mm, and HD scale bar represents 5mm.

**Figure S7.**
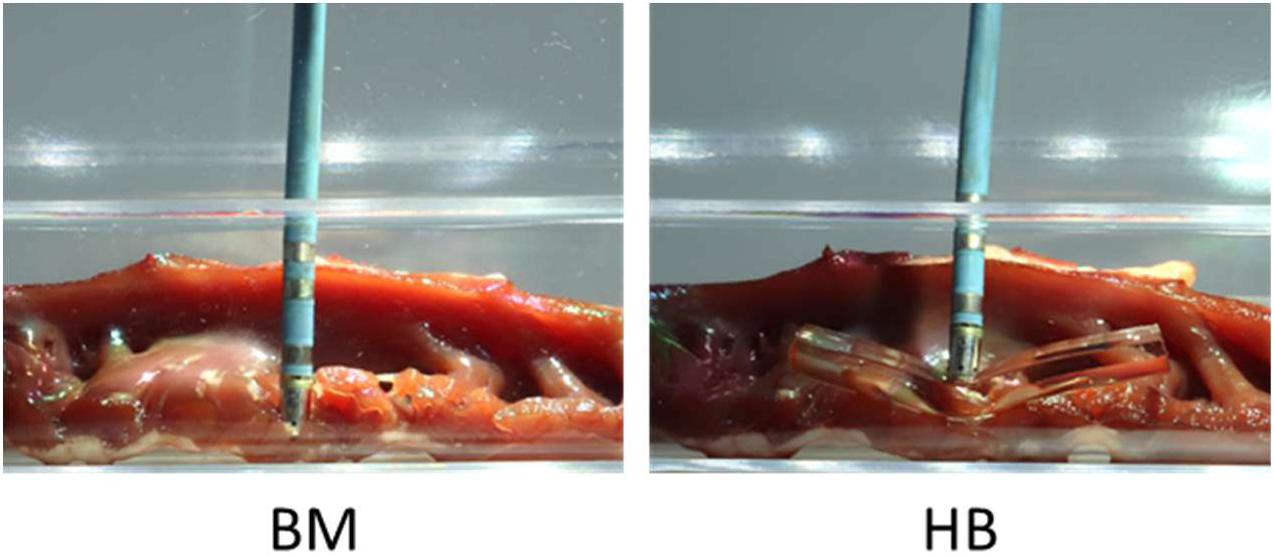
Conformability of the hydrogel electrode. Side-view photographs in the ablation tank of a standard bare-metal (BM) catheter electrode (left) and a hydrogel bar (HB) electrode (right) pressed against trabeculated cardiac tissue under the same applied load. The rigid BM tip contacts the tissue only at its apex, whereas the HB deforms to follow the tissue contour and contacts the surface along its length.

**Table S1.**
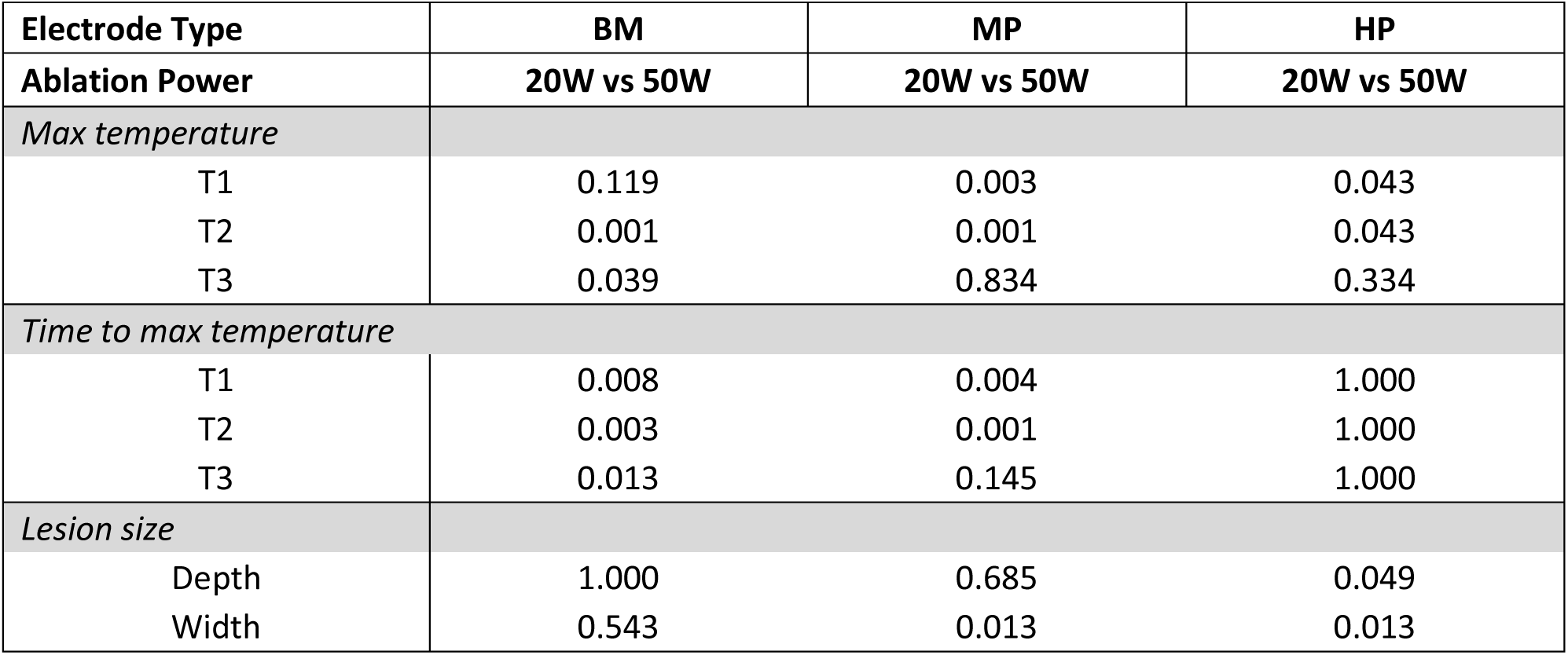
Comparison (p-values) of peak temperature, time to peak temperature, and lesion size measurements during ablations at 20W and 50W from BM, MP and HP. P-values are shown in each cell.

| Electrode Type | BM | MP | HP |
| --- | --- | --- | --- |
| Ablation Power | 20W vs 50W | 20W vs 50W | 20W vs 50W |
| *Max temperature* |  |  |  |
| T1 | 0.119 | 0.003 | 0.043 |
| T2 | 0.001 | 0.001 | 0.043 |
| T3 | 0.039 | 0.834 | 0.334 |
| *Time to max temperature* |  |  |  |
| T1 | 0.008 | 0.004 | 1.000 |
| T2 | 0.003 | 0.001 | 1.000 |
| T3 | 0.013 | 0.145 | 1.000 |
| *Lesion size* |  |  |  |
| Depth | 1.000 | 0.685 | 0.049 |
| Width | 0.543 | 0.013 | 0.013 |

**Table S2.**
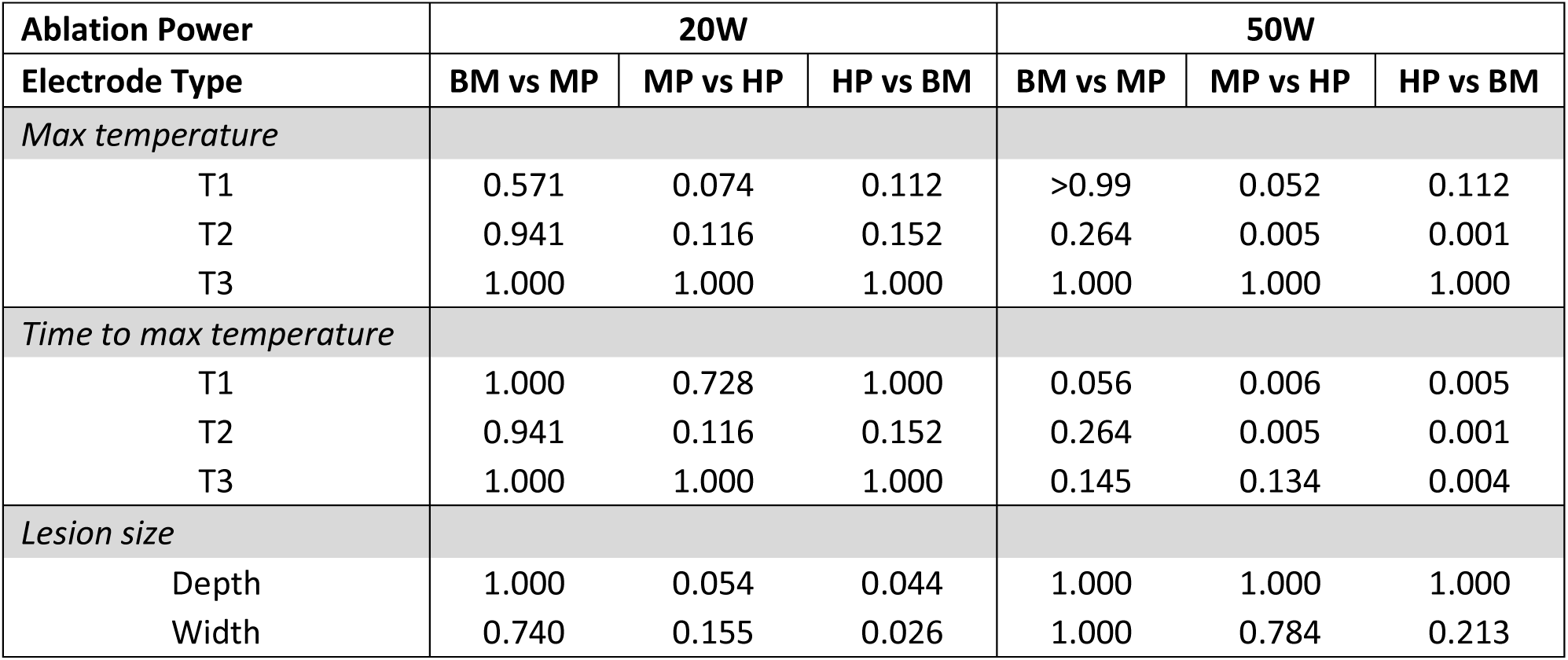
Comparison (p-values) of peak temperature, time to peak temperature, and lesion size measurements from BM, MP and HP when ablating with the same ablation parameter.

**Table S3.**
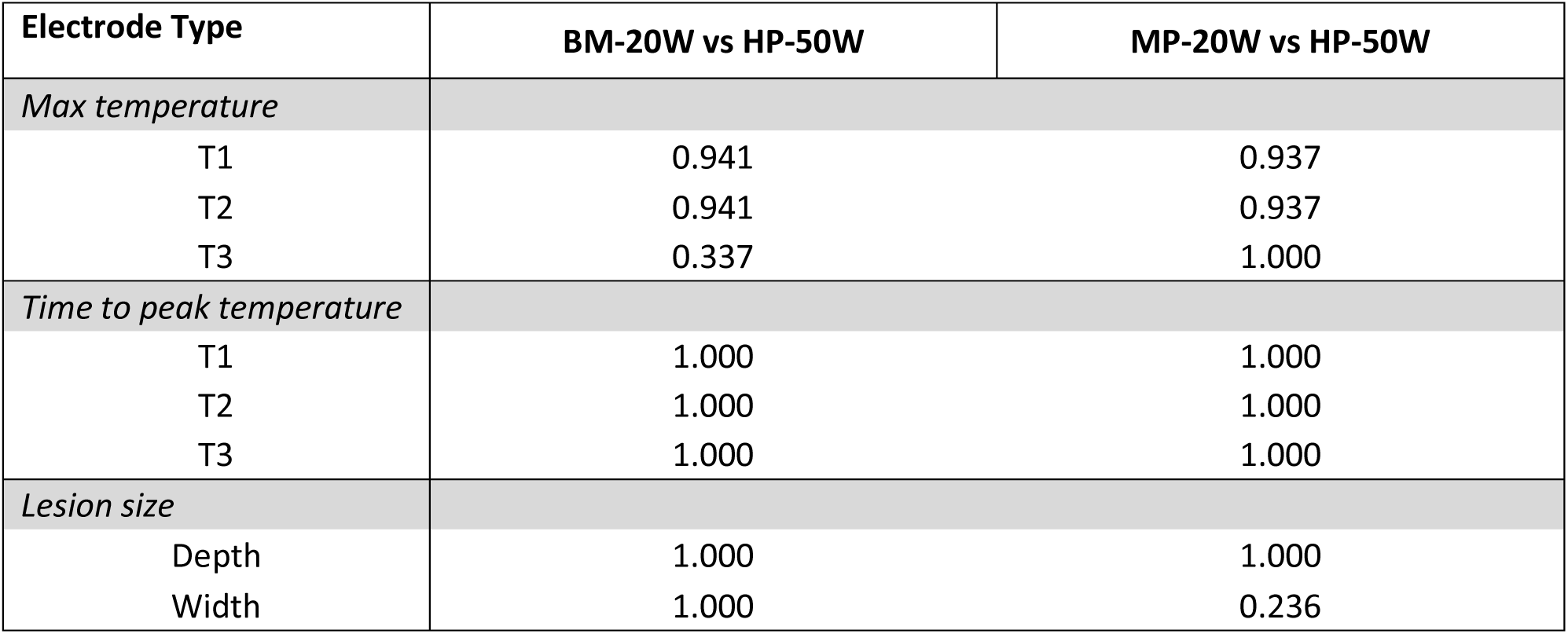
Comparison (p-values) of peak temperature of each probe, time to peak temperature, and lesion dimensions measured when ablating with BM and MP at 20W to HP at 50W.

